# Steady state haemolysis and cytoprotective protein levels in African children with sickle cell disease

**DOI:** 10.1101/2025.04.14.25325768

**Authors:** Emmanuel Alote Allotey, David Adedia, Kokou Hefoume Amegan-Aho, Enoch Aninagyei, Niiquaye Desmond Kwabena Quartey, Adjoa Agyemang Boakye, Verner Ndudiri Orish, Peter Wisdom Atadja, Lydia Mosi, Kwabena Obeng Duedu

## Abstract

Sub-Saharan Africa bears the highest burden of all Sickle Cell disease births worldwide. Chronic haemolysis in children with sickle cell disease (SCD) is known to cause multi-organ damage, increased risk of stroke, and cognitive impairment. This study sought to investigate haemolysis biomarkers in sickle cell disease (SCD) among sub-Saharan African children, aiming to improve individualized management through enhanced diagnostic and prognostic capabilities. Fifty children with SCD and 32 non-SCD children aged 2-17 years were evaluated using 5-part differential FBC and ELISA to profile haemolysis and haem cytoprotective proteins. Significantly elevated levels of bilirubin, free haemoglobin, haem oxygenase, ferritin, potassium ions, and AST activity in SCD participants was observed, while haptoglobin was significantly reduced. Hydroxyurea treatment was associated with increased haptoglobin and ferritin levels and decreased free haemoglobin and haem oxygenase activity. Male children with SCD exhibited higher haem oxygenase activity and free haemoglobin levels. A comparative analysis of machine learning algorithms revealed that the random forest model achieved 100% accuracy, sensitivity, specificity, predictive, and AUC values in classifying SCD. Direct bilirubin emerged as the most important classifier, followed by potassium, haptoglobin, free haemoglobin, haem oxygenase, total bilirubin, and ferritin. This research highlights the potential of machine learning-based classification using haemolysis biomarkers for improved SCD diagnosis and management. The findings from this sub-Saharan African cohort may have broader implications for SCD patient populations worldwide, potentially revolutionizing individualized treatment approaches and enhancing patient outcomes.

## Introduction

Sickle cell disease (SCD) is the most common genetic disease affecting the haemoglobin (Hb). Currently, an estimated 300,000 babies are born with SCD in the world annually, more than 75% of whom are born in sub-Saharan Africa (Piel et al., 2023). In Ghana, about 2% of newborns corresponding to approximately 15000 babies, are born with SCD annually (Ohene-Frempong et al., 2008; Segbefia et al., 2021). The most severe form is the homozygous HbS disease, which is found in 1% of Ghanaian babies born with SCD (Ohene-Frempong et al., 2008; Sundd et al., 2019). SCD is a debilitating and potentially fatal disease. Despite improvement of care, the risk of death due to SCD appears to increase with age in sub-Saharan Africa(Ranque et al., 2022). The clinical manifestations of SCD can be explained by an interplay of sickling and haemolysis (intravascular and extravascular), due to the abnormal haemoglobin which tend to polymerise under conditions causing low oxygen tension (Sundd et al., 2019). The sickling is primarily responsible for vaso-occlusion and eventually multiple organ damage while the haemolysis will cause anaemia, endothelial dysfunctions and Haemhaem-induced stimulation of inflammation which may worsen the vaso-occlusion(Sundd et al., 2019). Extravascular haemolysis is when red blood cells (RBC) containing polymerised Hb are ingested by the cells of the mononuclear phagocyte system, primarily in the spleen but also in the liver(Rapido, 2017). During intravascular haemolysis, free Hb and haem are released into the vasculature, potentially causing damage to the vasculature and exposing tissues through the effect of reactive oxygen species (ROS). Though individuals with only Hb S experience a higher rate of haemolysis as compared to those with Hb SC, in steady state, the intrinsic rate of haemolytic anaemia is relatively similar in the same individual (Mobark et al., 2020). The more prominent the haemolysis, the greater the risk of anaemia, vascular injury and end-organ damage, and this risk increases with age (Sundd et al., 2019).

In normal circumstances, cytoprotective proteins such as ham oxygenase-1 (HO-1) and haemopexin efficiently control these haem-related oxidative stress and cellular damage. Low levels of cytoprotective proteins in an individual with SCD increases the risk of complications while replenishing these protein levels may improve or even prevent sickle cell-related symptoms (Ashouri et al., 2021; Belcher et al., 2018).

Levels of markers of haemolysis and haem cytoprotective proteins are not routinely determined in our hospital laboratories owing largely to lack of logistics and reagents. Establishing the levels haemolysis and cytoprotective proteins is critical for effective clinical management of patients with SCD. The challenge however is that the reference levels are not representative of all populations and categories of patients. It is therefore important to establish baseline levels among patients that share common characteristics. This study aimed at determining the levels of haemolysis and cytoprotective proteins during steady state in African patients and their matched controls who do not have sickle cell disease.

## Methods

### Study Site, design and study participants

This case-control study was conducted among Ghanaian children aged 2-17 years following consent by parents or guardians. Child assent was also obtained from older children. Children that were studied were those with SCD (haemoglobin phenotypes S, SC, and SF) in steady state who have not been transfused blood or received any blood product within the previous 3 months and healthy non-SCD children were selected as comparative group. Participants who tested positive for malaria or had ongoing febrile illness or a history of febrile illness in the past week or had glucose-6-phospahte dehydrogenase deficiency were excluded from the study. The steady - state patient was identified based on the criteria employed by Akinola and colleagues. The study participants with SCD were recruited at the Ho Teaching Hospital whereas the comparative group were recruited from the Kodzobi Basic School (Control), both in the Volta Region of Ghana.

### Sample size determination

The number of samples to be analyzed were calculated based on the formula for determining the sample size for case-control studies published by Jaykaran Charan (2013).

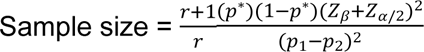

Where: *r* = ratio of control to cases (1:1); p* = average proportion of children less than 18 years with SCD = (proportion in cases + proportion in control)/2; Z_β_ = Standard normal variate for power of 80% (0.84), Z_a/2_ = Standard normal variate for level of significance (1.96); p1 – p2 = Effect size where p1 is proportion of anemia cases and p2 is proportion malaria in control. The proportion of Ghanaian children born with SCD was 2% (Ohene-Frimpong et al., 2008). Based on these parameters, the minimum sample size in each arm of the study was calculated to be 62.

### Sample collection and separation of sera

About 5 mL of venous blood was collected from each participant; 2 mL was dispensed into paediatric K_3_-EDTA tubes for full blood count assay and 3 mL was placed into a serum separator tube and allowed to clot. Serum was then collected from the serum separator tube and stored at −80^°^C until immunoassays were done.

### Laboratory procedures

#### Enumeration of formed cells and other associated parameters

Sysmex XN-550 (Kobe, Japan) was used to enumerate the formed cells and determine other calculated indices. The 5-part differential analyser works on the principle of laser beam multidimensional cell classification, flow cytometry for white cell differentiation, white and red cell estimation. Platelets were counted by optical and electrical impedance principles and haemoglobin concentration measured by cyanide-free colorimetric method. All other parameters were calculated automatically.

#### Determination of sickling status and Hb phenotype

Sickling status were determined using the sickling reagent 2% (W/V) sodium metabisulphite (Molar Mass = 190.107 g/mol) reagent. Briefly, about 2 μL of EDTA-anticoagulated blood was placed on a microscope slide, mixed with a drop (50 μL) of the sickling reagent and covered with a cover slip. Microscopy was done within 30 to 40 minutes of incubation at room temperature. The presence of sickled RBCs was confirmed by microscopy at x40 objective magnification (36). Hb genotype was determined using cellulose acetate alkaline electrophoresis (pH 8.6). Haemolysate was prepared by saline-washing of RBCs and lysing with 0.5% (v/v) Triton X-100 in 100 mg/L potassium cyanide. Electrophoresis was run on cellulose acetate membrane soaked in Tris-EDTA Borate (TEB) buffer (pH 8.4 - 8.6) by passing direct current delivering 350V at 50mA through the electrophoresis tank for 25 minutes. A positive control containing Hb C, S, F, and A were electrophoresed with each batch during phenotyping (37).

#### Enzyme immunoassays

The human haemoglobin (Cat. No. E88-134) and the human alpha-1-antitrypsin (Cat. No. E88-122) were done with reagents from Bethyl Laboratories Inc (Montgomery, TX 77356). The quantifications for human ferritin (product code ab108837) and human haptoglobin quantitation set (product code ab108856) were procured from Abcam PLC (Bristol, UK). The human Haem Oxygenase (HO)-1 (Cat. No. E0932Hu) quantitation was done with a kit from Bioassay Technology Laboratory (Shanghai, China).

#### Free plasma haemoglobin and human alpha-1-Antitrypsin quantification

The procedures for determination of free plasma haemoglobin and human alpha-1-antitrypsin levels were similar. For each assay, 100 μl of samples was added to designated wells (in duplicate). The plate was covered and incubated at room temperature (20-25 ° C) for an hour. The wells were then washed four times with 300 µL of the prepared wash buffer using an automated plate washer. Subsequently, 100 μl of anti-haemoglobin detection antibody or anti-alpha-1-antitrypsin detection anti-body was added to each well and the plate covered and incubated again at room temperature for an hour. The wells were washed four times after the incubation period. After washing, 100 μl of the kit specific HRP solution A was added to each well. The plate was covered and incubated at room temperature for thirty minutes. After thirty minutes of incubation, the wells were washed four times. After that, 100 μl of kit specific TMB substrate solution was then added to each well of the plate, incubated in the dark at room temperature for thirty minutes. Finally, the reaction was stopped by adding 100 μl of stop solution to each well. The absorbance was measured on a plate reader at 450nm with plate correction set at 630 nm. Each absorbance was converted to concentration using the standard curve that were prepared. The standard curves were prepared using the free plasma haemoglobin and human alpha-1-antitrypsin standard provided with kits. The absorbance for the standard were obtained as described for the sample testing.

#### Human Ferritin and haptoglobin quantitation

The procedures for human ferritin and haptoglobin quantitation were similar. In summary, 50 μL of serum was pipetted into labelled ferritin and haptoglobin wells and incubated for two hours. The wells were washed five times with 200 μL of 1X wash buffer manually and decanted. Subsequently, 50 μL of 1X biotinylated ferritin or biotinylated haptoglobin antibody was added to each well and incubated for one hour. Immediately after second washing, 50 μL of kit specific 1X SP conjugate was added to each well incubated for thirty minutes. After that, 50 μL of kit specific chromogen substrate was added to each well. The plate was incubated for 12 minutes or till the optimal blue colour density developed. The reactions were stopped with 50 μL of stop solution. The absorbance was read on a microplate reader at a wavelength of 450 nm (with plate correction at 630 nm) immediately. Concentrations of human ferritin and haptoglobin were obtained using standard curves prepared as shown by the manufacturer.

#### Human Haem Oxygenase 1 (HO-1) quantification

HO-1 quantification was done according to the manufacturer’s instructions. In brief, 40 μL of samples were added to each sample well followed by 10 μL anti-HO-1 antibody to sample wells, then 50 μL streptavidin-HRP to sample wells. The plate was incubated for sixty minutes at 37°C. After the incubation period, the wells were washed five times with 350 µL of the prepared wash buffer. The plate was then blotted onto paper towel. Later, 50 μL of substrate solution A was added to each well and then 50 μL of substrate solution B. The plate was covered and incubated for 10 minutes at 37°C in the dark. The reactions were stopped with 50 μL of stop solution. The absorbance of each well was determined immediately using a microplate reader set to 450 nm (plate corrected at 630 nm) within 10 minutes after adding the stop solution. Concentrations of HO-1 were obtained using standard curves prepared according to the manufacturer’s instruction.

### Statistical Analysis

All data and statistical analyses were performed using R version 4.3.1 and R-studio (2023.06.01). R packages such as ggpubr were used to draw the box plots comparing data between SCD and controls. Kruskal Wallis with multiple comparisons and Mann-Whitney tests were undertaken. The Kruskal Wallis test was followed by post-hoc analysis to compare between the Hb phenotypes (HbS, HbSC, HbSF) and controls. Analysis of variance and independent sample t-test was used to compare normally distributed datasets. Corrplot and Hmisc in R were used for the correlation plots. The correlation cut-offs were based on the rule of thumb for interpreting the size of a correlation coefficient as suggested by Hinkle and colleagues [30]. Briefly, correlations were analysed as follows: 0.90 to 1.00 (−0.90 to −1.00) - very high positive (negative) correlation; 0.70 to 0.90 (−0.70 to −0.90) - high positive (negative) correlation; 0.50 to 0.70 (−0.50 to −0.70) - moderate positive (negative) correlation; 0.30 to 0.50 (−0.30 to −0.50) - low positive (negative) correlation; and 0.00 to 0.30 (0.00 to −0.30) - negligible correlation. Boxplots were used to present comparative tests, while correlation plots were used to visualize correlations between the biomarkers. MASS, tidyverse, and ROCR in R were used for the discriminant analysis and ROC plot. Comparisons by heatmaps were generated using ComplexHeatmap in R. A test with p-value less than 0.05 was considered statistically significant. The biomarkers for haemolysis were also employed to classify participants as having SCD or not using some machine learning algorithms. The performance of the machine learning algorithms was compared considering measures such as accuracy, sensitivity, specificity, and the area under the curve (AUC) of the Receiver Operating Characteristic (ROC) curve.

## Results

### Participant demographic and Hb genotype distribution

Table 1 below describes the demographic and characteristics of the 82 study participants, including 50 (60.98%) cases and 32 (39.02%) controls. The mean age of the participants was 9.21 years (SD). Majority of the cases had homozygous Hb S while six out of the 32 controls were carriers of either HbS or HbC.

**Table 1:** Demographic, supplement and Hb phenotypes of the study participants.

|  | Characteristics | n (%) | Characteristics | n (%) |
| --- | --- | --- | --- | --- |
|  | **SCD Status** |  | **Multivitamins (SCD)** |  |
|  | Control | 32 (39.02) | Yes | 27 (54.00) |
|  | SCD | 50 (60.98) | No | 23 (46.00) |
|  | **Gender** |  | **Folic Acid (SCD)** |  |
|  |  |  | Yes | 40 |
|  | Male (SCD) | 25 (50.00) | (80.00) |  |
|  | Female (SCD) | 25 (50.00) | No | 10 (20.00) |
|  |  |  | **Zincovit (SCD)** |  |
|  | Male (Control) | 17 (53.13) | Yes | 20 (40.00) |
|  | Female (Control) | 15 (46.87) | No | 30 |
|  |  |  | (60.00) |  |
|  | **Hb Phenotype** |  | **Vit C (SCD)** |  |
| **Controls** | A | 26 (31.71) | Yes | 10 (20.00) |
|  | AC | 4 (4.88) | No | 40 (80.00) |
|  | AS | 2 (2.44) |  |  |
| **SCD** | S | 26 (31.71) | **Hydroxyurea** |  |
|  | SC | 9 (10.98) | Yes | 10 (20.00) |
|  | SF | 15 (18.29) | No | 40 (80.00) |

### Comparison of haematological profile between participants with SCD and healthy controls

Tables 2 compares parameters of the full blood count between participants with SCD and controls. All white cells parameters were significantly increased while red cells count was reduced among participants with SCD compared to the healthy controls. Platelets count was however similar between the two groups.

**Table 2:**
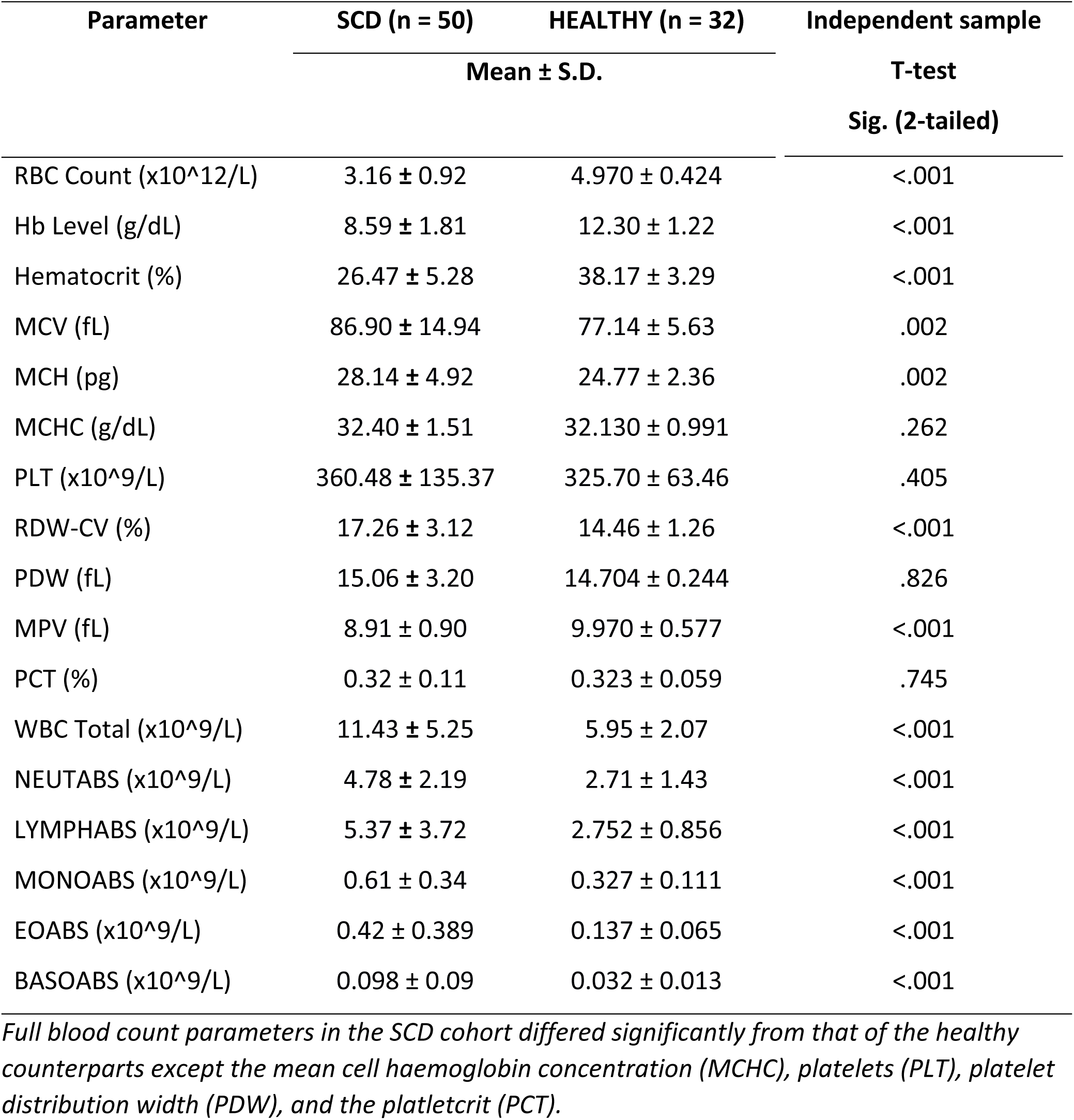
Comparison the FBC parameters between participants with SCD and healthy controls.

| Parameter | SCD (n = 50) | HEALTHY (n = 32) | Independent sample |
| --- | --- | --- | --- |
| | Mean ± S.D. | | T-test<br>Sig. (2-tailed) |
| RBC Count ($\times 10^{12}/L$) | 3.16 ± 0.92 | 4.970 ± 0.424 | <.001 |
| Hb Level (g/dL) | 8.59 ± 1.81 | 12.30 ± 1.22 | <.001 |
| Hematocrit (%) | 26.47 ± 5.28 | 38.17 ± 3.29 | <.001 |
| MCV (fL) | 86.90 ± 14.94 | 77.14 ± 5.63 | .002 |
| MCH (pg) | 28.14 ± 4.92 | 24.77 ± 2.36 | .002 |
| MCHC (g/dL) | 32.40 ± 1.51 | 32.130 ± 0.991 | .262 |
| PLT ($\times 10^9/L$) | 360.48 ± 135.37 | 325.70 ± 63.46 | .405 |
| RDW-CV (%) | 17.26 ± 3.12 | 14.46 ± 1.26 | <.001 |
| PDW (fL) | 15.06 ± 3.20 | 14.704 ± 0.244 | .826 |
| MPV (fL) | 8.91 ± 0.90 | 9.970 ± 0.577 | <.001 |
| PCT (%) | 0.32 ± 0.11 | 0.323 ± 0.059 | .745 |
| WBC Total ($\times 10^9/L$) | 11.43 ± 5.25 | 5.95 ± 2.07 | <.001 |
| NEUTABS ($\times 10^9/L$) | 4.78 ± 2.19 | 2.71 ± 1.43 | <.001 |
| LYMPHABS ($\times 10^9/L$) | 5.37 ± 3.72 | 2.752 ± 0.856 | <.001 |
| MONOABS ($\times 10^9/L$) | 0.61 ± 0.34 | 0.327 ± 0.111 | <.001 |
| EOABS ($\times 10^9/L$) | 0.42 ± 0.389 | 0.137 ± 0.065 | <.001 |
| BASOABS ($\times 10^9/L$) | 0.098 ± 0.09 | 0.032 ± 0.013 | <.001 |

*Full blood count parameters in the SCD cohort differed significantly from that of the healthy counterparts except the mean cell haemoglobin concentration (MCHC), platelets (PLT), platelet distribution width (PDW), and the plateletcrit (PCT).*

### Anaemia is a dominant phenotype in SCD and presents with varying degrees of severity

Anaemia status (Supplementary Table 1) was determined using cut-off limits as referenced by Janus & Moerschel (2010) and Wang (2016) (Janus & Moerschel, 2010; Wang, 2016). The proportion of anaemia was significantly higher among participants with SCD compared to controls (94.0% vs. 17.2%; p < .001). Similarly, the severity of anaemia was more pronounced among participants with SCD compared to controls with mild (26.9% vs. 12.5%), moderate (46.0% vs. 3.13%), and severe anaemia (18.0% vs. 0.0%)

### Levels of most haemolysis biomarkers are increased in SCD

The study compared direct markers of haemolysis such as K^+^, AST, Total, direct, and indirect bilirubin, and free Hb as well as haemolysis cytoprotective proteins such as haptoglobin, alpha 1 antitrypsin, ferritin, and haem oxygenase. The heat maps below (Figures 1 and 2) provide a quick view of the levels of the respective biomarkers of haemolysis measured among the study participants. Except for haptoglobin, all markers were increased in participants with SCD compared to controls (Figure 1) with marked increased among Hb S compared to Hb SC and SF (Figure 2).

**Figure 1:**
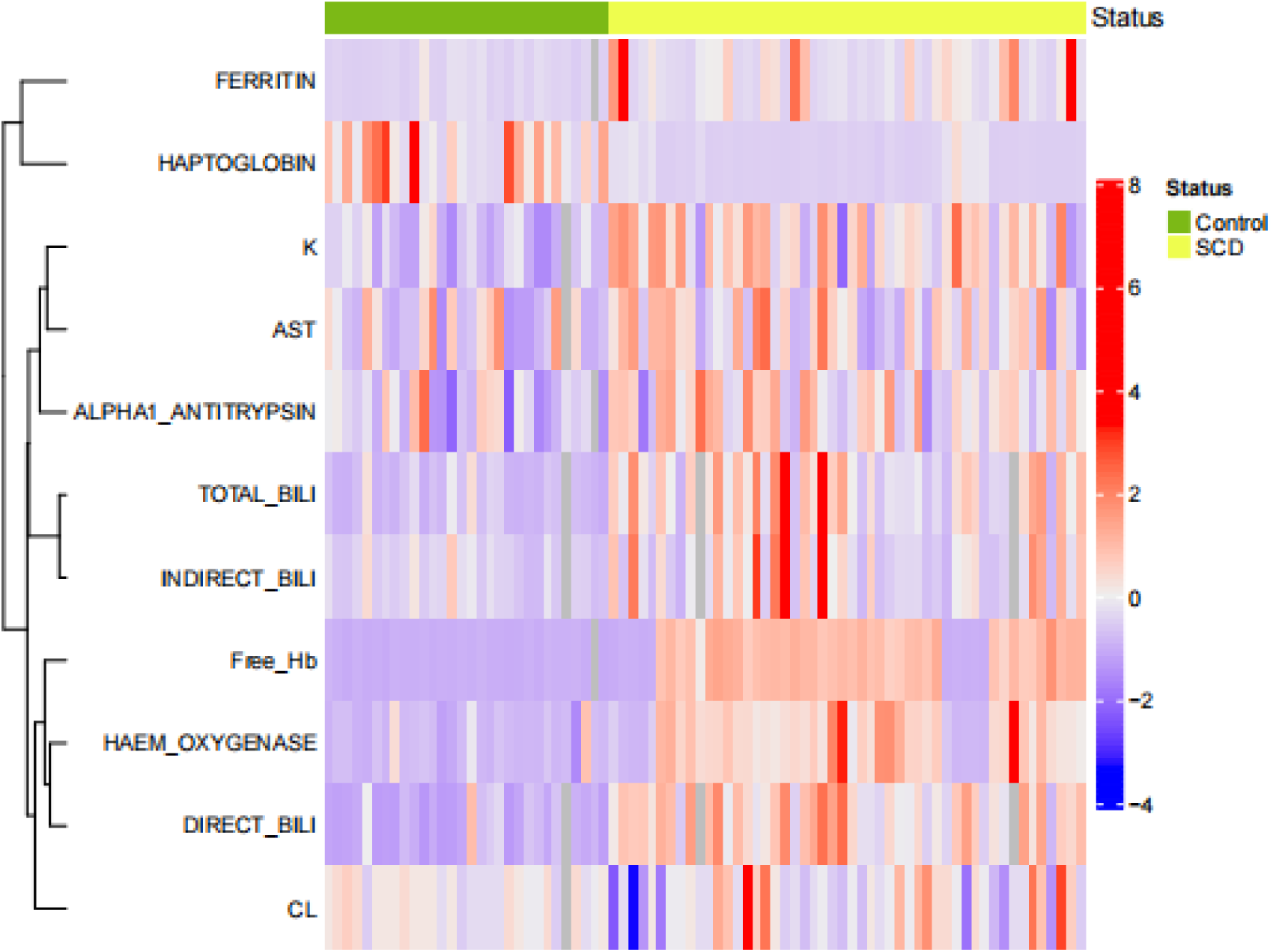
Comparison of markers of haemolysis between controls and participants with SCD using a heat map. Heat Map showing differences in levels of biomarkers of haemolysis between participants with SCD and controls. The red marks indicate an increase and the purple to blue indicate a decrease in the level of the biomarker.

**Figure 2:**
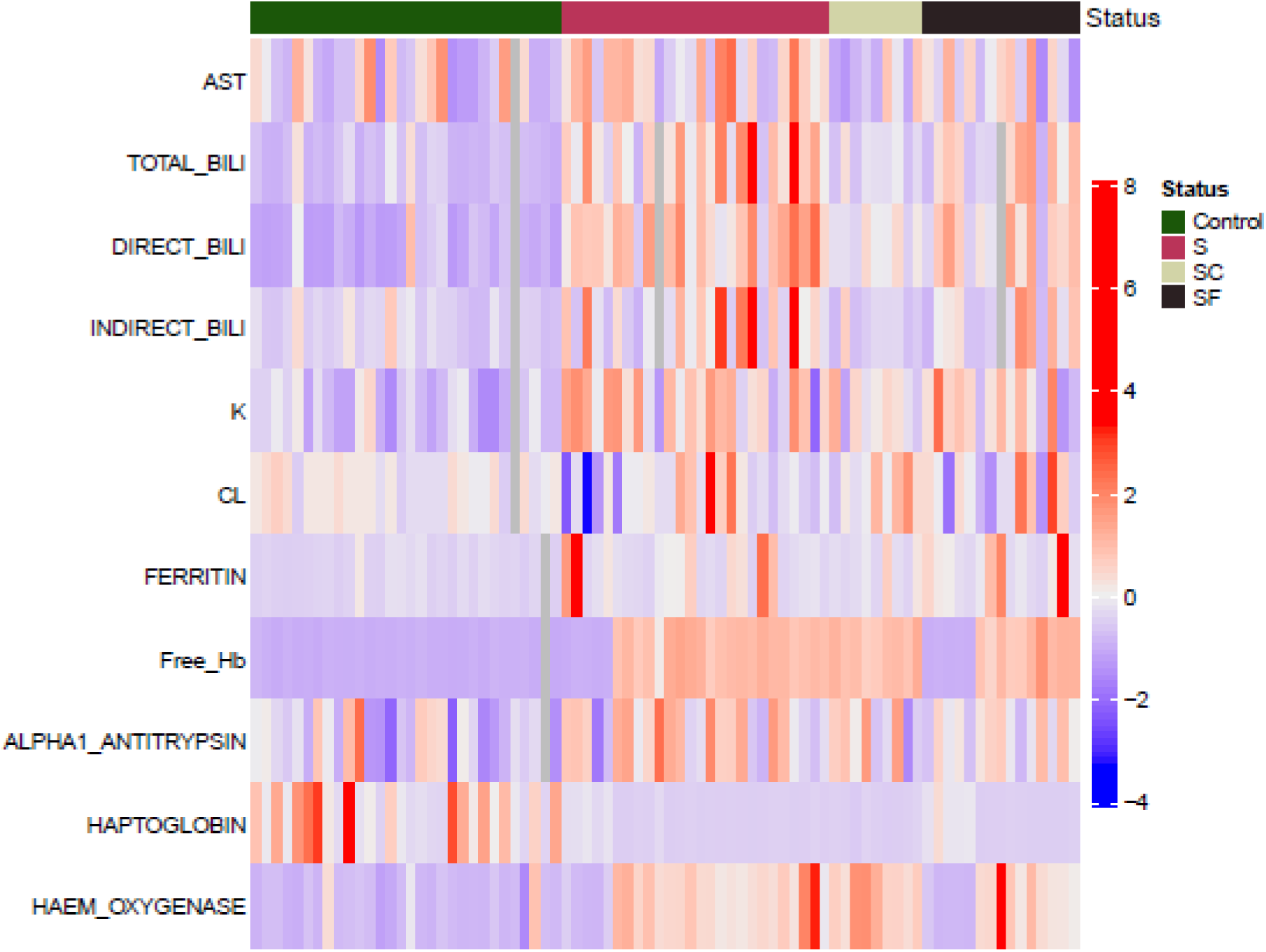
Comparison of markers of haemolysis between controls and SCD sub-phenotypes. Heat map shows differences in levels of biomarkers between controls and respective SCD sub-phenotypes. Most of the haemolysis biomarkers among the SCD sub-phenotypes are significantly increased. Haptoglobin levels however among the SCD sub-phenotypes appear significantly reduced.

The violin plots in Figure 3 further compare in detail the differences in the levels of the measured blood biomarkers between the controls and the SCD sub-phenotypes. Significant increases were observed in total bilirubin, direct bilirubin, and indirect bilirubin, as well as ferritin, potassium ions (K^+^), and free plasma Hb were observed in the SCD cohort compared to the control group (*p*<.05 for all). The activity of aspartate aminotransferase (AST), alpha-1-antitrypsin (AAT), and haem oxygenase were also significantly increased among the SCD phenotypes compared to the control group (*p*<.05 for all). However, plasma haptoglobin levels were significantly reduced among the SCD phenotypes compared to the controls. Haptoglobin levels were similar among the sub-phenotypes of SCD (*p*>.05). No significant difference was observed in chloride ions (Cl^-^)between the SCD and control groups (*p*>.05).

**Figure 3:**
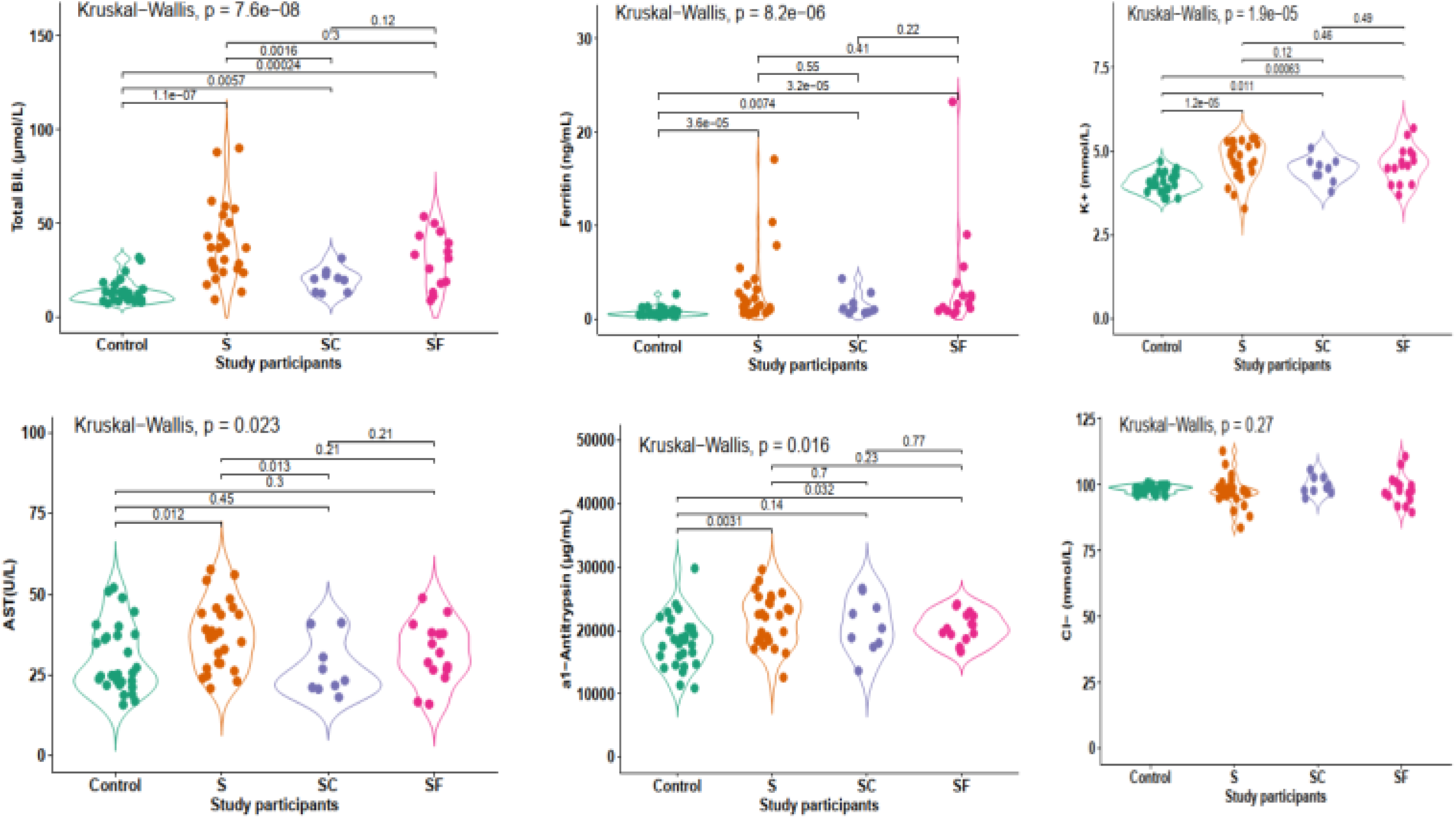
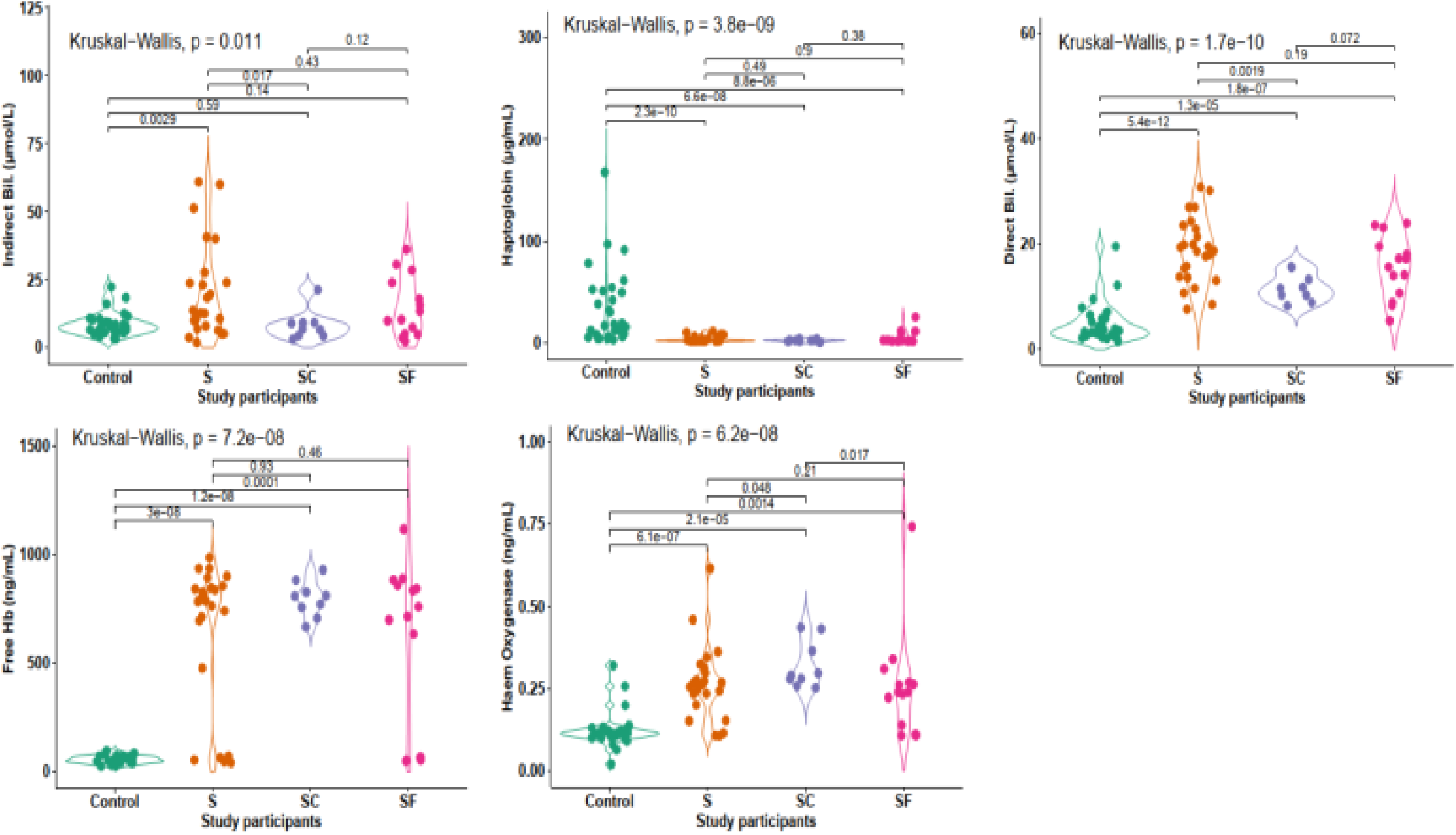
Violin plots comparing the statistical differences between controls and SCD sub-phenotypes. Levels of all the measured markers of haemolysis were significantly increased (*p*<.05) in the SCD sub-phenotypes compared to the controls. However, haptoglobin level was significantly reduced in the SCD sub-phenotypes. Chloride (Cl^-^)ions levels did not differ significantly between controls and SCD sub-phenotypes (*p*=.27)

For the following markers, no significant difference was observed between HbS and HbSF: Total, direct, and indirect bilirubin, ferritin, K+, free plasma Hb, haem oxygenase, haptoglobin, AST, and AAT (*p*>.05 for all).

The Total, direct, and indirect bilirubin, AAT, free plasma Hb, and AAT, and haem oxygenase activities were increased among the HbS phenotype compared to HbSC (*p*<.05). However, ferritin, K+, and AST activity was the same between HbS and HbSC phenotypes (*p*>.05).

All the measured markers of haemolysis except one were observed to be similar between the HbSC and HbSF phenotypes (*p*>.05). The exception was haem oxygenase activity which was found to be higher among the HbSC phenotypes (*p*<.05)

It was also observed that the Indirect bilirubin level and AST activity did not differ significantly between the controls, participants with HbSC, and HbSF (*p*>.05 for both).

### Haptoglobin and ferritin levels are increased while levels of free Hb and haem oxygenase activity are reduced in patients with SCD on hydroxyurea

Table 3 below compares the level of haemolysis biomarkers among patients with SCD on various medications and supplements for the management of the disease. Stratifying participants with SCD based on hydroxyurea use, the level of free Hb and haem oxygenase activity were significantly lower (*p*=.042 and *p*=.005 respectively) while the levels of ferritin and haptoglobin were significantly increased (*p*= .019 and *p*=.002) in the hydroxyurea group. Lower levels of free Hb were found among patients taking Zinc-containing multivitamins (Zincovit) (*p*=.025) while higher levels were found among patients taking Vitamin C (*p*=.031). However, no statistically significant difference (*p*>.05) was found in the level of any of the haemolysis biomarkers among patients taking non-Zinc-containing-multivitamins and folic acid.

**Table 3:** Comparison of levels of haemolysis biomarkers among SCD Sub-phenotypes based on use of hydroxyurea, multivitamins, folic acid, Zincovit, and Vitamin C.

| Biomarkers |  | Hydroxyurea | p-value | Multivitamins | p-value | Folic Acid | p-value | Zincovit | p-value | vitC | p-value |
| --- | --- | --- | --- | --- | --- | --- | --- | --- | --- | --- | --- |
| AST | Yes | 35±8.51 | .61 | 34.3±11.37 | .922 | 33.53±10.32 | .52 | 32.4±10.75 | .298 | 29.4±12.62 | .085 |
|  | No | 33.75±10.94 |  | 33.65±9.47 |  | 35.9±11.26 |  | 35.07±10.26 |  | 35.15±9.66 |  |
| TOTAL_BILI | Yes | 30.97±12.89 | .947 | 31.78±18.57 | .495 | 33.73±19.61 | .864 | 31.34±18.04 | .452 | 36.26±25.85 | .905 |
|  | No | 33.69±19.58 |  | 34.83±18.54 |  | 30.81±12.59 |  | 34.49±18.9 |  | 32.47±16.62 |  |
| DIRECT_BILI | Yes | 16.7±4.31 | .802 | 15.79±5.75 | .363 | 16.44±6.52 | .588 | 16.32±5.5 | .843 | 15.42±6.83 | .579 |
|  | No | 16.59±6.55 |  | 17.58±6.6 |  | 17.32±4.47 |  | 16.82±6.68 |  | 16.88±6.05 |  |
| INDIRECT_BILI | Yes | 14.27±10.84 | .947 | 16±14.62 | .521 | 17.28±15.42 | .822 | 15.02±14.11 | .53 | 20.84±19.53 | .721 |
|  | No | 17.1±15.43 |  | 17.25±14.93 |  | 13.48±10.6 |  | 17.68±15.12 |  | 15.59±13.37 |  |
| K<sup>+</sup> | Yes | 4.85±0.55 | .159 | 4.75±0.54 | .198 | 4.66±0.55 | .752 | 4.71±0.54 | .796 | 4.51±0.45 | .35 |
|  | No | 4.6±0.52 |  | 4.53±0.5 |  | 4.59±0.49 |  | 4.61±0.53 |  | 4.68±0.55 |  |
| CL<sup>-</sup> | Yes | 96.09±5.76 | .567 | 97.34±5.29 | .487 | 97.72±5 | .283 | 98.43±5.9 | .96 | 97.7±4.22 | .354 |
|  | No | 98.74±5.18 |  | 99.24±5.34 |  | 100.18±6.48 |  | 98.07±5.04 |  | 98.34±5.63 |  |
| Ferritin | Yes | 6.61±7.69 | .019 | 3.03±3.6 | .546 | 3.15±4.67 | .174 | 2.86±3.91 | .635 | 3.68±3.59 | .467 |
|  | No | 2.09±2.1 |  | 2.96±4.91 |  | 2.4±1.17 |  | 3.09±4.46 |  | 2.83±4.37 |  |
| FreeHb | Yes | 309.63±400.86 | .042 | 614.25±345.65 | .202 | 648.59±333.25 | .753 | 567.13±355.76 | .025 | 795.34±271.33 | .031 |
|  | No | 748.78±227.1 |  | 715.77±283.34 |  | 710.37±266.4 |  | 723.5±281.76 |  | 627.35±324.71 |  |
| A1AT | Yes | 20382.72±3512.06 | .561 | 21581.14±3308.46 | .355 | 20669.43±3417.08 | .069 | 21215.44±4285.34 | .782 | 21744.18±3269.19 | .513 |
|  | No | 21407.96±3744.22 |  | 20758.91±4118.58 |  | 23336.83±4134.8 |  | 21194.56±3307.49 |  | 21067.6±3810.64 |  |
| Haptoglobin | Yes | 9.56±6.89 | .002 | 5.49±5.18 | .066 | 4.86±4.63 | .734 | 5.66±5.64 | .406 | 3.51±2.91 | .308 |
|  | No | 3.52±2.3 |  | 3.83±2.97 |  | 4.19±3.13 |  | 4.1±3.18 |  | 5.03±4.62 |  |
| Haem_Oxygenase | Yes | 0.21±0.2 | .005 | 0.26±0.13 | .284 | 0.27±0.13 | .78 | 0.24±0.09 | .216 | 0.3±0.17 | .818 |
|  | No | 0.29±0.09 |  | 0.28±0.11 |  | 0.26±0.06 |  | 0.29±0.13 |  | 0.26±0.1 |  |

### Haem oxygenase activity and free Hb level are increased among male children with SCD

In Table 4, the level of the haemolysis biomarkers was compared among males and females with and without SCD. No significant differences were found between male and female control samples. However, males with SCD presented with significantly higher activity for haem oxygenase (*p*=.025) and increased levels of free Hb (*p*=.026) compared to the females. No statistically significant difference (*p*>.05) in the other haemolysis biomarkers was found between males and females with SCD.

**Table 4:**
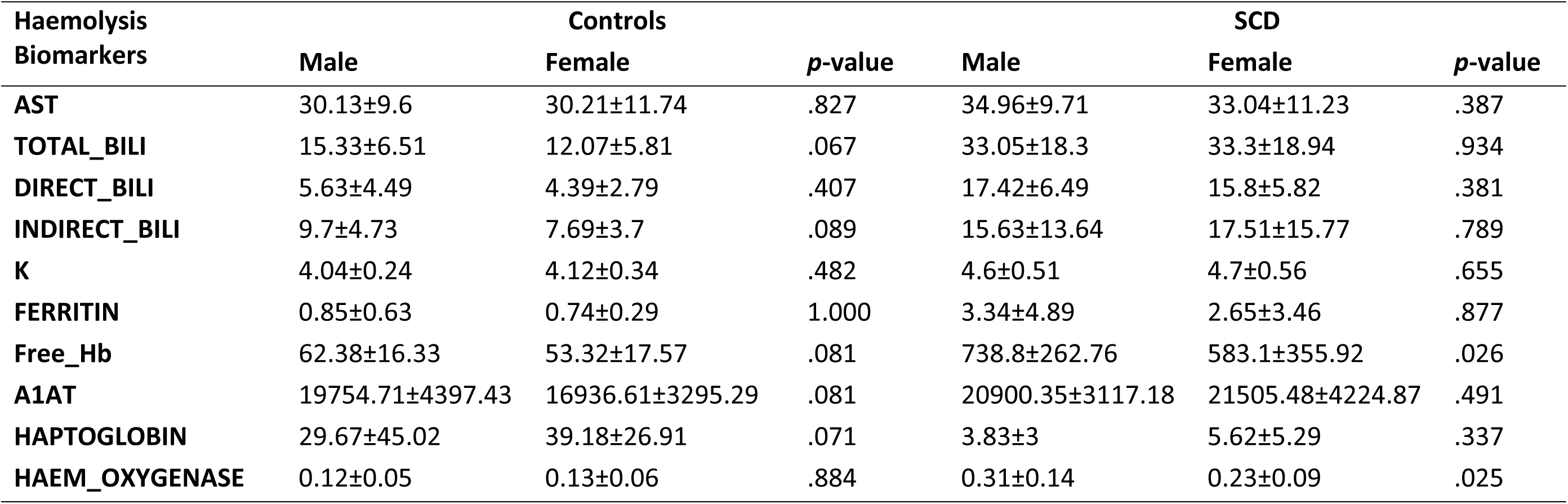
Comparing the level of haemolysis biomarkers in males and females with and without SCD.

| Haemolysis Biomarkers | Controls |  |  | SCD |  |  |
| --- | --- | --- | --- | --- | --- | --- |
|  | Male | Female | *p*-value | Male | Female | *p*-value |
| **AST** | 30.13±9.6 | 30.21±11.74 | .827 | 34.96±9.71 | 33.04±11.23 | .387 |
| **TOTAL_BILI** | 15.33±6.51 | 12.07±5.81 | .067 | 33.05±18.3 | 33.3±18.94 | .934 |
| **DIRECT_BILI** | 5.63±4.49 | 4.39±2.79 | .407 | 17.42±6.49 | 15.8±5.82 | .381 |
| **INDIRECT_BILI** | 9.7±4.73 | 7.69±3.7 | .089 | 15.63±13.64 | 17.51±15.77 | .789 |
| **K** | 4.04±0.24 | 4.12±0.34 | .482 | 4.6±0.51 | 4.7±0.56 | .655 |
| **FERRITIN** | 0.85±0.63 | 0.74±0.29 | 1.000 | 3.34±4.89 | 2.65±3.46 | .877 |
| **Free_Hb** | 62.38±16.33 | 53.32±17.57 | .081 | 738.8±262.76 | 583.1±355.92 | .026 |
| **A1AT** | 19754.71±4397.43 | 16936.61±3295.29 | .081 | 20900.35±3117.18 | 21505.48±4224.87 | .491 |
| **HAPTOGLOBIN** | 29.67±45.02 | 39.18±26.91 | .071 | 3.83±3 | 5.62±5.29 | .337 |
| **HAEM_OXYGENASE** | 0.12±0.05 | 0.13±0.06 | .884 | 0.31±0.14 | 0.23±0.09 | .025 |

### Correlations between haemolysis biomarkers and full blood count parameters among study groups

The following correlations were observed between markers of haemolysis and variables in the full blood count profiles of the participants with SCD (figure 4a):

**Figure 4:**
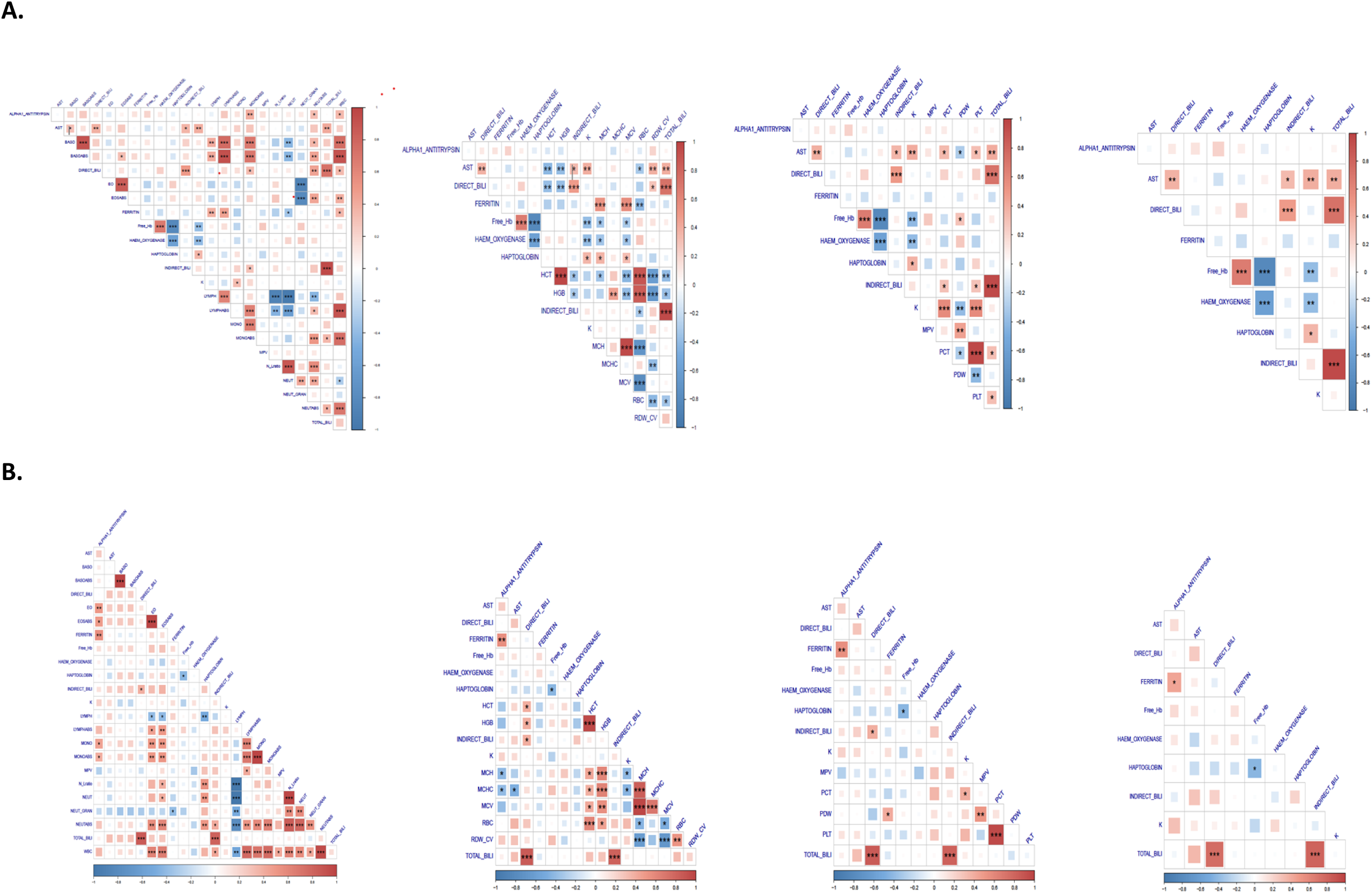
Correlations showing the relationship between the haem cytoprotective proteins and the full blood count variables. **A** represents the correlations for the SCD sub-phenotypes and **B** represents correlations for the control group.

Alpha-1-antitrypsin activity correlated positively with the absolute monocyte, and neutrophil counts, as well as the total WBC counts (*p*<.05 for all).

AST activity positively correlated with PCT, PLT count, relative basophil counts (BASO), the direct, indirect, and total bilirubin, K^+^, and RDW-CV (*p*<.05 for all). A negative correlation was however observed between AST activity and HCT, HGB, RBC count, and PDW (*p*<.05 for all)

Positive correlations were observed between direct bilirubin (DIRECT_BILI), and total bilirubin (TOTAL_BILI), Indirect bilirubin (INDIRECT_BILI), absolute monocyte and neutrophil counts, total WBC, and RDW-CV (*p*<.05 for all). On the contrary, DIRECT_BILI negatively correlated with and HCT, and HGB (*p*<.05 for all).

The INDIRECT_BILI levels in the participants with SCD also correlated positively with TOTAL_BILI levels, absolute monocyte counts, PCT, and PLT (*p*<.05). The INDIRECT_BILI levels, however, correlated negatively with the RBC count (*p*<.05).

The TOTAL_BILI levels positively correlated with the absolute neutrophil and monocyte counts, PCT, and PLT count (*p*<.05).

Ferritin levels positively correlated with the total WBC count, and absolute lymphocyte count as well as the percentage lymphocyte count. Additionally, there was a positive correlation between ferritin levels, and the MCV and MCH (*p*<.05 for all). There was, however, a negative correlation between the ferritin level, and the percentage neutrophil count, and RBC count (*p*<.05).

Free haemoglobin (Free Hb) levels positively with the PDW and haem oxygenase levels (*p*<.05). However, negative correlations were seen between the free Hb level, and haptoglobin, and potassium ions (K^+^) level. The free Hb level also correlated negatively with the MCH and MCV (*p*<.05).

Haem oxygenase activity positively correlated with K^+^ level but correlated negatively with the MCV and MCH (*p*<.05 for all).

Haptoglobin levels correlated positively with K^+^ level, and negatively with free Hb and haem oxygenase activity (*p*<.05 for all)

Potassium (K^+^) level also correlated positively with percentage monocyte count, the PCT and PLT count but correlated negatively with the PDW (*p*<.05 for all).

### Correlations between markers of haemolysis and full blood count variables of the participants without SCD (controls)

Figure 4B shows all the correlations as observed among the control group.

Among the controls, Alpha-1-antitrypsin activity correlated positively with the absolute monocyte count (*p*<.05). However, the relationship was stronger among the SCD sub-phenotypes compared to the controls. Other positive correlations were also observed between alpha-1-antitrypsin levels and percentage eosinophil, absolute eosinophil, and percentage monocyte counts (*p*<.05 for all) among the controls. These relationships were not, however, significant among the SCD sub-phenotypes (*p*>.05 for all). The alpha-1-antitrypsin activity level also correlated positively with ferritin levels among the controls (*p*<.05). This relationship though observed among the SCD group, was not statistically significant (*p*<.05).

A negative correlation was observed between AST activity and the MCHC (*p*<.05). This relationship was not observed among the participants with SCD.

Positive correlations were observed between DIRECT_BILI and TOTAL_BILI, INDIRECT_BILI levels, HGB and HCT (*p*<.05 for all) among the control group. A negative correlation was, on the contrary, observed between DIRECT_BILI and HCT, and HGB (*p*<.05 for all) in the SCD group. Comparatively, there was a stronger positive correlation between direct bilirubin and indirect bilirubin among the SCD sub-phenotypes compared to the control group.

The INDIRECT_BILI levels in the control group correlated positively with TOTAL_BILI levels and the strength of the correlation was similar to what was observed among the SCD sub-phenotypes (*p*<.05). The INDIRECT_BILI levels in the control also correlated positively with the absolute neutrophil count, and total WBC count (*p*<.05 for both). However, this relationship was not observed among the SCD sub-phenotypes.

The TOTAL_BILI levels positively correlated with DIRECT_BILI and INDIRECT_BILI levels (*p*<.05 for both) The strengths of these correlations were similar to what was observed among the SCD sub-phenotypes. No significant correlation was found between the TOTAL_BILI levels and absolute neutrophil and monocyte counts, PCT, and PLT count as observed among the SCD sub-phenotypes (*p*>.05 for all).

Ferritin levels positively correlated with alpha-1-antitrypsin activity among the controls (*p*<.05) This relationship although present among the SCD sub-phenotypes, was not statistically significant (*p*>.05). Compared to the SCD sub-phenotypes, there was no significant correlation observed between ferritin and absolute lymphocyte count, and percentage lymphocyte count among the controls. Additionally, there was no significant positive correlation between ferritin levels and the MCV and MCH (*p*>.05 for all). There was also no significant correlation between the ferritin level and the percentage neutrophil count, and RBC counts (*p*>.05 for both).

A negative correlation was observed between Free Hb and haptoglobin levels among the controls (*p*<.05). However, the strength of the relationship was lower compared to the strength of the relationship among the SCD sub-phenotypes. None of the other correlations observed among the SCD sub-phenotypes concerning free Hb was found to be significant among the controls (*p*>. 05 for all).

Haptoglobin levels correlated negatively with free Hb (*p*<.05) among the controls as reported earlier. However, the other correlations reported earlier among the SCD sub-phenotypes between haptoglobin and K^+^ level, and haem oxygenase activity (*p*<.05 for all) were not statistically significant among the controls (*p*>.05 for both).

Potassium (K^+^) level also correlated positively with the PCT among the controls (*p*<.05). However, the relationship among the SCD group was stronger compared to what was observed among the controls. A negative correlation was observed between K^+^ level and the MCH, and MCHC (*p*<.05 for both). This correlation was not significant among the SCD sub-phenotypes. The correlations between K^+^ level and PCT, and PLT count observed among the SCD phenotypes were not significant among the controls (*p*>.05 for both).

### Cluster analysis

The cluster analysis correctly classified individuals into two groups (SCD and controls) individuals, which was supported by silhouette and gap statistic methods. The values of the biomarkers classified most sickle cell patients together in cluster 1 and healthy controls with a few with SCD in cluster 2 (Figure 5). This shows that the values of biomarkers used can usually classify individuals as having SCD or controls. A few SCD cases that were classified among controls may be those simultaneously on HU, Zincovit, and Folic acid.

**Figure 5:**
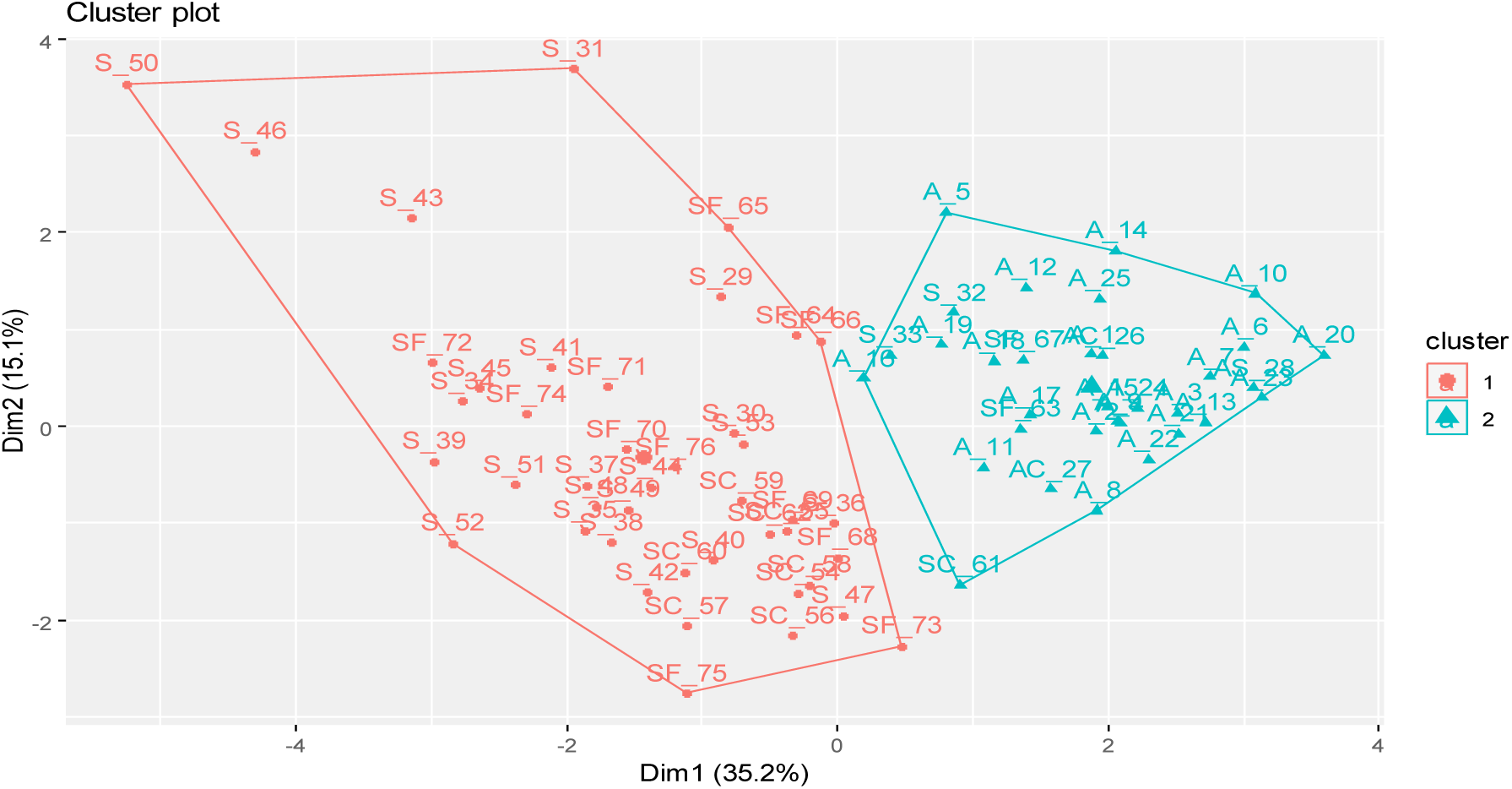
Cluster biplots and dendrogram of biomarkers.

### Classification of individuals as SCD patients using machine learning algorithms

Using biomarkers to classify and predict individuals with SCD, a comparative analysis of machine learning algorithms, such as generalized linear model (GLM), random forest (RF), Decision tree, and k nearest neighbour, was undertaken, which reported model assessment measures in Table 5. The random forest model outperformed other machine learning models, by reporting the best measures, including, accuracy, sensitivity, specificity, and the area under the curve (AUC) of the Receiver Operating Characteristic (ROC) curve. Figure 6 below shows the importance plot of the biomarkers for the random forest model in predicting SCD. The direct bilirubin was the best classifier, followed by K+, haptoglobin, free haemoglobin, haem oxygenase, total bilirubin, and ferritin.

**Figure 6:**
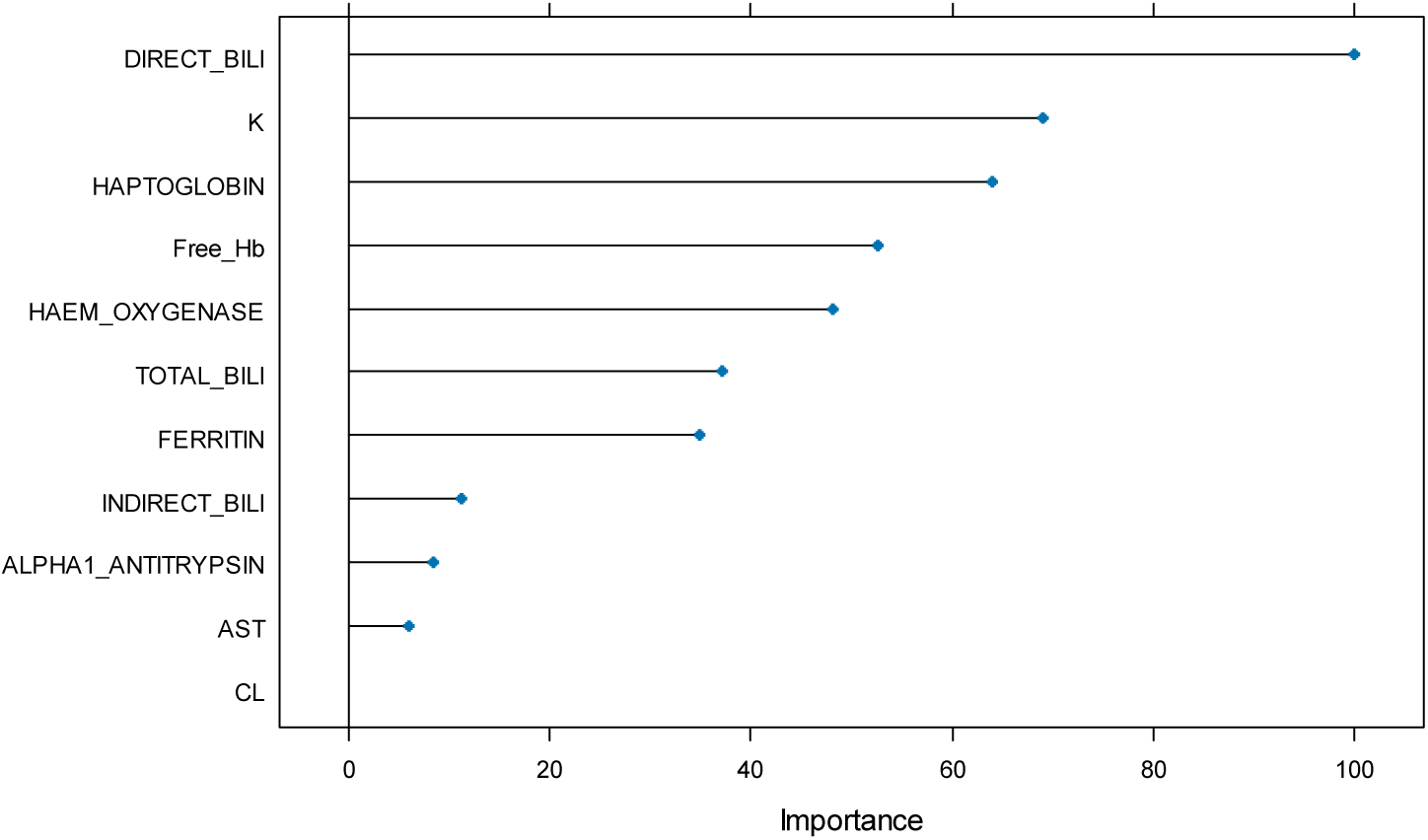
SCD prediction importance of biomarkers.

**Table 5:** Classification algorithms for predicting SCD.

| Algorithms | Accuracy | Sensitivity | Specificity | Positive predictive value | Negative predictive value | AUC |
| --- | --- | --- | --- | --- | --- | --- |
| GLM | 0.96 | 0.93 | 1.00 | 1.00 | 0.90 | 0.95 |
| RF | 1.00 | 1.00 | 1.00 | 1.00 | 1.00 | 1.00 |
| Decision tree | 0.96 | 0.93 | 1.00 | 1.00 | 0.90 | 0.97 |
| KNN | 0.71 | 0.93 | 0.33 | 0.70 | 0.75 | 0.84 |

## Discussion

The study investigated haemolysis biomarkers and their relationship with full blood count parameters among children with SCD in the Volta region of Ghana. The results describe the demographic distribution, supplements, and medications of the study participants. Comparative analysis of the various measured haemolysis biomarkers shows significantly increased levels among patients with SCD compared to controls. Levels of haptoglobin, a cytoprotective protein, were however markedly reduced in SCD. Hydroxyurea therapy and the use of some supplements appeared to modulate levels of some of the haemolysis biomarkers. Gender-based comparison reported increased haem oxygenase activity and free haemoglobin in males with SCD. Correlation analysis also showed relationships between some of the measured biomarkers and full blood count parameters. The biomarkers effectively classified individuals into respective study groups as shown by cluster analysis. High prediction accuracy was also reported by machine learning algorithms like random forest in classifying individuals as having SCD or not.

The age range for this study falls within the common paediatric age range for most SCD studies (Boadu et al., 2018; Egesa et al., 2022). As in other reports, there was no gender predominance observed among the participants with SCD (Awaitey et al., 2020; Boadu et al., 2018; Ceglie et al., 2019). Over 50% of the participants reported the use of multivitamins and folate which likely represents a high level of supplementation. Folate supplementation helps prevent folate deficiency and macrocytosis in SCD (Dixit et al., 2018; Francis et al., 2022). However, evidence for a definitive positive effect on clinical outcomes remains equivocal (Dixit et al., 2018). The low number of participants on hydroxyurea likely represents the low prescription rates in most African settings (Dexter & McGann, 2023; Ofakunrin et al., 2021). Hydroxyurea remains the only approved disease-modifying therapy for SCD and its use in eligible patients needs to improve (Dexter & McGann, 2023).

The heat map provides a good visual overview of the levels of the various haemolysis biomarkers. The significantly increased levels of AST, bilirubin (total, direct and indirect), potassium ions, and free haemoglobin represent increased haemolysis. Hyperbilirubinemia results from accelerated breakdown of haemoglobin and reflects the degree of haemolysis (Singh et al., 2024). The Unconjugated or indirect bilirubin binds to albumin and is not excreted in urine while conjugated or direct bilirubin is water soluble and excreted. Conjugated hyperbilirubinemia as occurs in SCD patients reflects saturation of the conjugation mechanism by excessive haemolysis (Shah et al., 2017). The increased serum AST also reflects haemolysis as well as hepatic congestion from sickling (Nsiah et al., 2011). The reduced haptoglobin levels demonstrate increased consumption resulting from its role in scavenging free haemoglobin during haemolysis (Smith & McCulloh, 2015). Free haemoglobin readily oxidizes to methaemoglobin which releases redox active haem/iron that cause oxidative damage to cells (Belcher et al., 2014). Haptoglobin forms a stable complex with free haemoglobin preventing its oxidative and renal effects (Quaye, 2008). The increased haem oxygenase activity likely functions to degrade the excess free haemoglobin as a homeostatic mechanism against haemoglobin toxicity (Belcher et al., 2014). The comparable chloride ion levels agree with other reports indicating it is not typically altered by haemolysis in SCD (Antwi-Boasiako et al., 2019).

The increased ferritin levels would reflect increased iron stores resulting from chronic haemolysis and blood transfusion therapy in SCD. However, none of the participants with SCD were being chronically transfused. The increased ferritin levels in these patients may be reflecting the sub-clinical inflammation that characterizes the steady state of SCD and would be expected to be higher during crisis.. (Van Beers et al., 2015). The increased alpha-1 antitrypsin likely functions as an antioxidant and anti-inflammatory agent with a high binding affinity to free Haem, thus reducing endothelial cell injury caused by free haem (Madyaningrana et al., 2021). Its upregulation possibly mitigates oxidative damage from haemolysis.

The violin plots further clarify the statistically significant differences between the SCD subgroups and controls regarding each biomarker. This aids in appreciating differences in degree of haemolysis between the respective SCD phenotypes. As reported in studies, patients with the HbS phenotypes manifest the most severe clinical disease due to the higher degree of haemolysis and sickling. The HbSC phenotype represents a milder form of the disease (Rees et al., 2022).

Hydroxyurea therapy significantly reduced free haemoglobin and haem oxygenase activity indicating its role in reducing haemolysis. Hydroxyurea induces HbF synthesis which inhibits HbS polymerization (Agrawal et al., 2014). Decreased sickling leads to reduced haemolysis. The increased haptoglobin and reduced ferritin levels in hydroxyurea-treated patients likely results from reduced haemolysis and inflammation respectively (Pule et al., 2015). Though needing confirmation in larger studies, the modulation of some biomarkers by supplements likely reflects their role as adjunct therapy in SCD (Crouch et al., 2018). Ascorbic acid possesses antioxidant properties that may reduce oxidative stress and haemolysis (Gęgotek & Skrzydlewska, 2022). Zinc plays vital roles in antioxidant pathways, immune function and growth (Prasad & Bao, 2019). It further protects against lipid peroxidation in RBCs and consequently stabilizes biomembranes and biostructures thus protecting against oxidative stress. Its modulation of free haemoglobin agrees with its role in reducing sickling and haemolysis (Thaisa de Oliveira et al., 2022). Low Zinc level among patients with SCD has been documented.

The higher haem oxygenase activity and free haemoglobin in males possibly indicates increased haemolysis. The Lower HbF levels reported among males with SCD compared with females may have contributed to this observation. Similar observations have been reported regarding haemolysis markers between genders (Raslan et al., 2018). Males often present more severe manifestations of SCD. The exact mechanism for the gender difference remains unclear though androgen and oestrogen effects have been postulated in patients who have reached puberty (Ceglie et al., 2019).

Correlation analysis aids in assessing the interrelationships between the measured biomarkers and full blood count indices. This helps identify associated trends between haemolysis markers, anaemia severity and aberrant haematological changes in SCD. As expected, total and direct bilirubin levels inversely correlated with haemoglobin levels since both reflect haemolysis (Hansen et al., 2020)

The positive correlations of AST with platelet and white cell counts likely result from its role as a marker of haemolysis and inflammation in SCD (Nsiah et al., 2011). Inflammation stimulates leucocytosis and thrombocytosis in patients with SCD (Conran & Belcher, 2018). The associated increase in platelet and white cell counts may be suggestive of a hepatic steatosis which in most cases is benign and asymptomatic in SCD (Chao et al., 2022). The positive correlations between potassium ions and platelet indices also likely represent an inflammatory effect (Conran & Belcher, 2018). Though needing further studies, the negative correlation between potassium ions and PDW possibly suggests high potassium levels impair platelet formation and reactivity. PDW measures variation in platelet sizes and higher values occur with increased generation of larger and more reactive platelets (Vagdatli et al., 2010).

The positive correlation between ferritin and lymphocyte count may relate to increased inflammation in SCD which also stimulates neutrophil and monocyte proliferation (Zhang et al., 2016). The positive correlation between ferritin and MCV/MCH agrees with the role of iron deficiency in causing microcytic hypochromic anaemia (Mohammed et al., 2020; Rusch et al., 2023). Hence, increase in ferritin levels within the normal range in SCD would suggestively keep the MCV AND MCH levels within their respective normal ranges. This further suggests that a reduced or increased MCH and MCV may be an indicator of poorly controlled SCD. However, an increased MCV is a marker of compliance to hydroxyurea treatment (Gordeuk & Gordeuk, 2018).

Free haemoglobin positively correlating with PDW suggests it possibly stimulates production of larger platelets (Annarapu et al., 2021). Free haemoglobin is also known to disrupt the redox balance which activates platelets (Schaer et al., 2013). The negative correlation between free haemoglobin and haptoglobin agrees with the role of haptoglobin in reducing and scavenging free haemoglobin respectively (Gbotosho et al., 2021; Schaer et al., 2013).

Overall, fewer significant correlations existed between the biomarkers and full blood count parameters among controls compared to the SCD participants. The significant correlations in the control group also mostly showed weaker strengths. This suggests the haemolysis process and aberrant compensatory mechanisms in SCD forge pathophysiologic relationships between the biomarkers and full blood count indices.

Cluster analysis aids in assessing the capability of measured biomarkers to delineate individuals into their respective study groups (Lynch & DeGruttola, 2022). As shown, the biomarkers effectively clustered most of the SCD patients into one dominant cluster separate from the controls. This indicates the composite biomarker profile distinguishes sickle cell patients from controls. The random forest algorithm also showed excellent performance in classifying individuals as having SCD with perfect accuracy. Variable importance ranking also identified the most discriminatory biomarkers as direct bilirubin, potassium ions, and haptoglobin among others. This further demonstrates the utility of the haemolysis biomarkers as excellent discriminant tools for identifying persons with a haemolytic disease.

The study has some notable limitations. The lack of data on HbF levels and α-globin gene status limited assessment of their modulation of the haemolysis biomarkers. Elevated HbF inhibits HbS polymerization and reduces haemolysis in SCD patients (M. H. Steinberg et al., 2014) Co-inheritance of α-thalassemia also ameliorates the degree of haemolysis in SCD (Rees et al., 2022; Rumaney et al., 2014). Data on these modifiers would have aided in establishing their role in modulating haemolysis biomarkers.

The inclusion of only paediatric subjects also limits generalization to adult populations with SCD. Significant differences exist between paediatric and adult patients regarding clinical severity, haemolysis profiles, and mortality risk (Heeney et al., 2014). Data on hydroxyurea use was also limited to establish its definitive effects. Assessing serial changes before and after hydroxyurea initiation would better describe its role.

Additionally, evaluating intravascular haemolysis alone provides limited insight into the overall complex pathophysiology of SCD. This is based on the premise that intravascular haemolysis contributes to only one-third of haemolysis in SCD (Steinberg et al., 2009) Complementing the biomarkers with markers of extravascular haemolysis like erythrocyte phosphatidylserine exposure would have offered more composite assessment (Boas et al., 1998). Providing data on chronic organ damage like albuminuria would have also aided in correlating haemolysis severity with end-organ damage. Establishing genetic modifiers of haemolysis like UGT1A1 genotypes would have further enriched the findings as polymorphisms of *UGT1A1* promoter has been associated with bilirubin levels and gall stones in patients of African descent with SCD (Hamad et al., 2013; Milton et al., 2012).

### Study Limitations

Some limitations should be considered when interpreting the study results. As a cross-sectional analysis, it provides a snapshot of haematological and haemolysis markers that may fluctuate over time. Longitudinal monitoring could better characterize dynamic changes with disease progression, hydroxyurea treatment, acute complications, etc. The sample size, although appropriate for common statistical tests, remains relatively small. Subgroup analyses by SCD genotype and hydroxyurea use had limited power to detect smaller differences between groups.

Additionally, the inclusion of only paediatric patients prevented comparisons to adult SCD populations. Further analyses across wider age spectrums could provide fuller understanding of haemolysis throughout the lifespan of the Ghanaian patient with SCD.

In summary, this study provides important confirmatory evidence regarding haematological abnormalities and haemolysis biomarkers in SCD patients. It reinforces the central role of chronic intravascular haemolysis and haemoglobinopathy-induced inflammation and oxidative stress in the pathophysiology of SCD. Several findings merit further research, including elucidating mechanisms for increased haemolysis in males, standardizing the pattern of haemolysis markers across different genotypes, and clarifying hydroxyurea’s effects on haemolysis over time

## Conclusion & Recommendations

Overall, the study provides useful data on haemolysis profiles of a paediatric SCD population in Ghana. Significant relationships identified between haemolysis biomarkers and full blood count indices also offer insight into associated haematological aberrations in SCD. Elucidating the role of hydroxyurea and supplements in modulating haemolysis also guides optimizing treatment. Findings from the study provide an impetus for larger follow-up studies to further describe haemolysis profiles and relationships to clinical outcomes in the population. Efforts at early identification of patients with increased haemolysis can guide individualized risk stratification and treatment. Ultimately, further dissection of haemolysis phenotypes and genotypes in SCD can guide individualized treatment approaches. Therapies tailored to each patient’s degree of anaemia, haemolytic rate, inflammatory state, and end-organ dysfunction may offer optimal outcomes. As highlighted in this study, haemolysis biomarkers provide a window into the underlying disease process and response to interventions. Ongoing research can illuminate their evolving role in prognosis, monitoring, and personalized management of SCD.

## Data Availability

All data produced in the present study are available upon reasonable request to the authors

## Statements & Declarations

### Author Contributions

Conceptualization: EAA, KHAA, PWA, KOD; Data curation: EAA, NDKQ; Formal Analysis: EAA, DA, KOD; Funding acquisition and resources: EAA, PWA, KOD; Methodology and Investigation: EAA, DA, KHAA, EA, NDKQ, AAB, VNO; Visualization: EAA, DA, KOD; Writing – original draft: EAA, KHAA, DA, EA, VNO, KOD; Writing – review & editing: All Authors; Supervision: PWA, LM, KOD

### Data Availability

Relevant data has been reported in the manuscript. Any other where needed will be made available upon a reasonable request from the corresponding author. Contact email: Contact

### Ethics Approval

This study was performed in line with the principles of the Declaration of Helsinki. The study was approved by the Research Ethics Committee of the University of Health and Allied Sciences, Ho with protocol reference numbers UHAS-REC A.3 [1] 18-19 and UHAS-REC A.12 [42] 20-21. Permission was sought from the Research Directorate of the Ho Teaching Hospital (HTH/RPPME/19/1) and the Adaklu District Directorate of the Ghana Education Service (GES/VR/ADEO/48/VOL.1/31).

### Consent to Participate

For children below age of 16 informed consent was obtained from their parents. In addition, child ascent was obtained for children 5 years and above. For children above age of 16 informed consent was obtained from them as well as their parents.

### Consent to Publish

Not Applicable

## Acknowledgements

We wish to acknowledge the contribution of the staff of the sickle cell clinic at the Ho Teaching Hospital, Mr. Daniel Mensah and the dietetics team, Mr. Idan Baah Banson, and the Laboratory Team of the Ho Teaching Hospital for offering various forms of support during recruitment of the patients.

